# Place and underlying cause of death during the COVID19 pandemic: retrospective cohort study of 3.5 million deaths in England and Wales, 2014 to 2020

**DOI:** 10.1101/2020.08.12.20173302

**Authors:** Jianhua Wu, Marion Malfam, Mamas Mamas, Muhammad Rashid, Evangelos Kontopantelis, John E Deanfield, Mark A de Belder, Chris P Gale

## Abstract

**Background:** The COVID-19 pandemic has resulted in a high death toll. We aimed to describe the place and cause of death during the COVID-19 pandemic.

**Methods:** This national death registry included all adult (aged ≥18 years) deaths in England and Wales between 1^st^ January 2014 and 30^th^ June 2020. Analyses were based upon ICD-10 codes corresponding to the underlying cause of death as stated on the Medical Certificate of Cause of Death. Daily deaths during COVID-19 pandemic were compared against the expected daily deaths estimated using Farrington surveillance algorithm for daily historical data between 2014 and 2020, by place and cause of death.

**Findings:** Between 2^nd^ March and 30^th^ June 2020, there was an excess mortality of 57,860 (a proportional increase of 35%) compared with the expected deaths, of which 50,603 (86.2%) were COVID-19 related. Almost half the excess deaths occurred in care homes (25,611 deaths) where deaths were 55% higher than expected. One fifth of the excess deaths occurred in hospital (15,938 deaths; a proportional increase of 21%) with the remainder occurring at home (16,190 deaths; a proportional increase of 39%). At home, only 14% of 16,190 excess deaths were related to COVID-19, with 5,963 deaths due to cancer and 2,485 deaths due to cardiac disease, very few of which involved COVID-19. In care homes or hospices, 61% of the 25,611 excess deaths were related to COVID-19, 5,539 of which were due to respiratory disease and most of these (4,315 deaths) involved COVID-19. In hospital, there were 16,174 fewer deaths than expected which did not involve COVID-19, and there were 4,088 fewer deaths due to cancer and 1,398 fewer deaths due to cardiac disease than expected.

**Interpretation:** The COVID-19 pandemic has resulted in a substantial increase in the absolute numbers of deaths occurring at home and care homes. There was a huge burden of excess deaths occurring in care homes, which were poorly characterised, and were likely to be, at least in part, the result of undiagnosed COVID-19. There was a smaller but important and ongoing excess in deaths at home, particularly from cancer and cardiac disease, which suggests avoidance of hospital care for non-COVID-19 conditions.

**Funding:** The study is unfunded.

## Introduction

Globally, as of August 6^th^ 2020, coronavirus disease (COVID-19) caused by severe acute respiratory syndrome coronavirus 2 (SARS-CoV-2) accounted for 702,642 deaths.^1^ In the UK, this has been evidenced by an abrupt increase in the number of deaths above that expected for the historical average.^2 3^ However, the basis for this excess mortality is poorly defined with limited about the causes of death during the pandemic. This is important because whilst the SARS-CoV-2 virus is known to result in an acute respiratory syndrome for whom the highest risk of death is among the elderly and those with pre-existing medical conditions,^4 5^ but also people may have died from other causes due restructuring of medical services during this period or avoidance of health care settings

Moreover, we and others have reported a dramatic decline in admissions to hospitals with medical emergencies.^6–11^ Consequent to delays to seeking help for life-threatening illnesses many deaths are likely to have occurred in the community. Equally, there has been an increase in the number of deaths among those living in care homes.^12–14^ Here, the vulnerability of residents to infection as well as changes to health behaviour may hay played a role in their demise. Should there have been a displacement in the place of death as a result of the pandemic, then lessons may be learnt to be better prepared in case of a second increase in COVID-19 cases.^15^

Thus, a systematic characterisation of the cause and place of death associated with the COVID-19 pandemic and how this changed compared to pre-pandemic era is necessary and may offer insights into the susceptibility of the public to the virus as well as the impact of health and public guidance aimed at reducing the spread of the virus. We report the underlying causes of all adult deaths during the COVID-19 pandemic in England and Wales, the location of deaths (e.g. hospitals, home or care homes) and their relation to the COVID-19 infection. This information is vital for the understanding of healthcare policy during the emergence from lockdown and to assist Governments around the world reorganise healthcare services now that incident rates of COVID-19 are in decline and social isolation policies are relaxed.

## Methods

### Data

The analytical cohort included all certified and registered deaths in England and Wales ≥18 years of age, between 1^st^ January 2014 and 30^th^ June 2020 recorded in the Civil Registration Deaths Data of the Office for National Statistics (ONS) of England and Wales.^16^

### Deaths

The primary analysis was based upon the ICD-10 code corresponding to the underlying cause of death registered, as stated on the medical certificate of cause of death (MCCD). The MCCD is completed by the doctor who attended the deceased during their last illness within 5 days unless there is to be a coroner’s post-mortem or an inquest. Underlying causes of death were then categorised as detailed in the Office for National Statistics (ONS) short list for causes of death,^17^ with additional aggregation of causes for cancer, cardiac and respiratory pathologies (Supplement table 1). ICD-10 codes ‘U071’ (confirmed) and ‘U072’ (suspected) listed in any position on the MCCD were used to identify whether a death involved COVID-19 infection. For the purposes of this investigation, the ICD-10 code corresponding to the underlying cause of death was counted only once per deceased. Pre-existing conditions or other diseases that contributed to, but did not directly lead to death were excluded from the analyses. We found that about one in ten MCCDs reported COVID-19 as the underlying cause of death, and for such cases we used the pathology named as directly leading to death to select the underlying cause, which is also the approach taken by the ONS.^17^ The place of death as recorded on the MCCD was classified as home, care home or hospice, and hospital.

### Statistical analyses

Baseline characteristics were described using numbers and percentages for categorical data. Data were stratified by COVID-19 status (suspected or confirmed COVID-19 recorded, not mentioned), age band (<50, 50-59, 60-69, 70-79, 80+ years)), sex and place of death. The number of daily deaths was presented using a 7-day simple moving average (the mean number of daily deaths for that day and the preceding 6 days) from 1^st^ February 2020 up to and including 30^th^ June 2020, adjusted for seasonality.

The expected daily deaths from 1^st^ February 2020 up to and including 30^th^ June 2020 were estimated using Farrington surveillance algorithm for daily historical data between 2014 and 2020.^18^ The algorithm uses overdispersed Poisson generalised linear models with cubic spline terms to model trends in counts of daily death, accounting for seasonality. The number of non-COVID-19 deaths each day from 1^st^ February 2020 were subtracted from the estimated expected daily deaths in the same time period to create a zero historical baseline. Deaths above this baseline may be interpreted as excess mortality, which were calculated as the difference between the observed daily deaths and the expected daily deaths. The proportion of excess deaths was estimated by dividing the excess mortality by the sum of the expected deaths between 2^nd^ March 2020 and 30^th^ June 2020. To compare the impact on mortality from COVID-19 pandemic and flu epidemic, information about influenza and pneumonia (ICD-10 code: J09-J18) was extracted for the two months either side of the date of the peak death rate each year between 2015 and 2020. The averaged daily deaths over 6 six years in the ‘influenza season’ were compared with the averaged daily deaths in the trough period(two months before and after 1^st^ July each year). All tests were two-sided and statistical significance considered as *P*<0.05. Statistical analyses were performed in R version 4.0.0.

### Ethical approval

Ethical approval was not required as this study used fully anonymised routinely collected civil registration deaths data. The data analysis was conducted through remote access to NHS Digital Data Science Server.

## Results

Between 1^st^ January 2014 and 30^th^ June 2020, there were 3,451,538 deaths from all-causes among adults in England and Wales, of which 224,615 (6.5%) deaths occurred after 2^nd^ March 2020. While hospital remained the most frequent place of death, compared with prior to 2^nd^ March 2020 there were proportionally fewer deaths in hospital (41% vs. 48%) and more in care homes and hospices (33% vs. 29%), with similar proportions at home (26% vs 24%) (Table 1).

**Table 1.** Deaths before and after 2nd March 2020, by COVID-19 status *The numbers do not add up to the total deaths due to missingness (1.9%).

|  | Deaths before 2<sup>nd</sup><br>March 2020 | Non-COVID-19<br>related deaths<br>after<br>2<sup>nd</sup> March 2020 | COVID-19<br>related<br>deaths after<br>2<sup>nd</sup> March 2020 | Deaths after<br>2<sup>nd</sup> March 2020 |
| --- | --- | --- | --- | --- |
| Total | n = 3,226,923 | n = 174,012 | n = 50,603 | n = 224,615 |
| **Sex** |  |  |  |  |
| Male | 1,584,621 (49.1) | 85,594 (49.2) | 27,863 (55.1) | 113,457 (50.5) |
| Female | 1,642,302 (50.9) | 88,418 (50.8) | 22,740 (44.9) | 111,158 (49.5) |
| **Age category (years)** |  |  |  |  |
| 18-49 | 136,823 ( 4.2) | 6,444 ( 3.7) | 995 ( 2.0) | 7,439 ( 3.3) |
| 50-59 | 183,594 ( 5.7) | 9,934 ( 5.7) | 2,312 ( 4.6) | 12,246 ( 5.5) |
| 60-69 | 374,105 (11.6) | 18,932 (10.9) | 4,883 ( 9.6) | 23,815 (10.6) |
| 70-79 | 708,387 (22.0) | 38,886 (22.3) | 11,376 (22.5) | 50,262 (22.4) |
| 80+ | 1,824,014 (56.5) | 99,816 (57.4) | 31,037 (61.3) | 130,853 (58.3) |
| **Region** |  |  |  |  |
| North East | 146,146 ( 5.3) | 9,056 ( 5.2) | 2,832 ( 5.6) | 11,888 ( 5.3) |
| North West | 374,758 (13.6) | 23,490 (13.5) | 7,741 (15.3) | 31,231 (13.9) |
| Yorkshire and The Humber | 272,521 ( 9.9) | 17,214 ( 9.9) | 4,691 ( 9.3) | 21,905 ( 9.8) |
| East Midlands | 219,614 ( 8.0) | 14,050 ( 8.1) | 3,574 ( 7.1) | 17,624 ( 7.8) |
| West Midlands | 289,234 (10.5) | 18,692 (10.7) | 5,848 (11.6) | 24,540 (10.9) |
| East of England | 291,819 (10.6) | 19,027 (10.9) | 4,902 ( 9.7) | 23,929 (10.7) |
| London | 268,237 ( 9.7) | 16,851 ( 9.7) | 8,606 (17.0) | 25,457 (11.3) |
| South East | 425,229 (15.4) | 26,591 (15.3) | 7,124 (14.1) | 33,715 (15.0) |
| South West | 295,718 (10.7) | 18,778 (10.8) | 2,879 ( 5.7) | 21,657 ( 9.6) |
| Wales | 169,288 ( 6.2) | 10,260 ( 5.9) | 2,404 ( 4.8) | 12,664 ( 5.6) |
| **Place of death\*** |  |  |  |  |
| Home | 760,173 (24.0) | 55,324 (32.5) | 2,334 ( 4.6) | 57,658 (26.2) |
| Care home or hospice | 900,691 (28.5) | 56,179 (33.0) | 15,966 (31.7) | 72,145 (32.7) |
| Hospital | 1,503,836 (47.5) | 58,546 (34.4) | 32,112 (63.7) | 90,658 (41.1) |
| **Underlying cause of death** |  |  |  |  |
| Malignant neoplasms | 896,515 (27.8) | 46,932 (27.0) | 1,127 ( 2.2) | 48,059 (21.4) |
| Cardiac diseases | 497,556 (15.4) | 26,230 (15.1) | 1,050 ( 2.1) | 27,280 (12.1) |
| Respiratory diseases | 451,324 (14.0) | 20,769 (11.9) | 19,861 (39.2) | 40,630 (18.1) |
| Dementia and Alzheimer disease | 393,405 (12.2) | 26,168 (15.0) | 1,036 ( 2.0) | 27,204 (12.1) |
| Cerebrovascular diseases | 199,578 ( 6.2) | 9,734 ( 5.6) | 441 ( 0.9) | 10,175 ( 4.5) |
| Symptoms, signs and ill-defined conditions | 76,228 ( 2.4) | 6,351 ( 3.6) | 2,718 ( 5.4) | 9,069 ( 4.0) |
| Diseases of the urinary system | 53,264 ( 1.7) | 2,882 ( 1.7) | 280 ( 0.6) | 3,162 ( 1.4) |
| Cirrhosis and other diseases of liver | 50,404 ( 1.6) | 2,820 ( 1.6) | 116 ( 0.2) | 2,936 ( 1.3) |
| Diabetes | 36,619 ( 1.1) | 2,631 ( 1.5) | 164 ( 0.3) | 2,795 ( 1.2) |
| Parkinson's disease | 36,295 ( 1.1) | 2,515 ( 1.4) | 91 ( 0.2) | 2,606 ( 1.2) |
| Other cause of diseases | 535,735 (16.6) | 26,980 (15.5) | 1,784 ( 3.5) | 28,764 (12.8) |
| COVID only | - | - | 21,935 (43.3) | 21,935 (9.8) |

\*The numbers do not add up to the total deaths due to missingness or unknown place of death (1.9%).

### Excess deaths after 2^nd^ March 2020

The peak in deaths during the COVID-19 pandemic was much greater than for any of the influenza seasonal peaks in the years between 2015 and 2020 (Figure 1). Following the first COVID-19 death on the 2^nd^ March 2020, there was an excess mortality of 57,860 (a proportional increase of 35%) compared with the expected daily deaths estimated using Farrington surveillance algorithm for daily historical data between 2014 and 2020 (Figure 2, Table 2). The number of excess deaths was higher for men than women (29,956; a proportional increase of 36% vs. 27,839; a proportional increase of 33%), and was the highest among people aged over 80 years (37,244; a proportional increase of 40%) (Table 2). London had the largest absolute number of excess deaths (9,001 deaths; a proportional increase of 55%). Almost half the excess deaths occurred in care homes and hospices (25,611 deaths) where deaths were 55% higher than expected. One quarter of the excess deaths occurred in hospital (15,938 deaths; a proportional increase of 24%) with the remainder occurring at home (16,190 deaths; a proportional increase of 39%) (Table 2).

**Figure 1.**
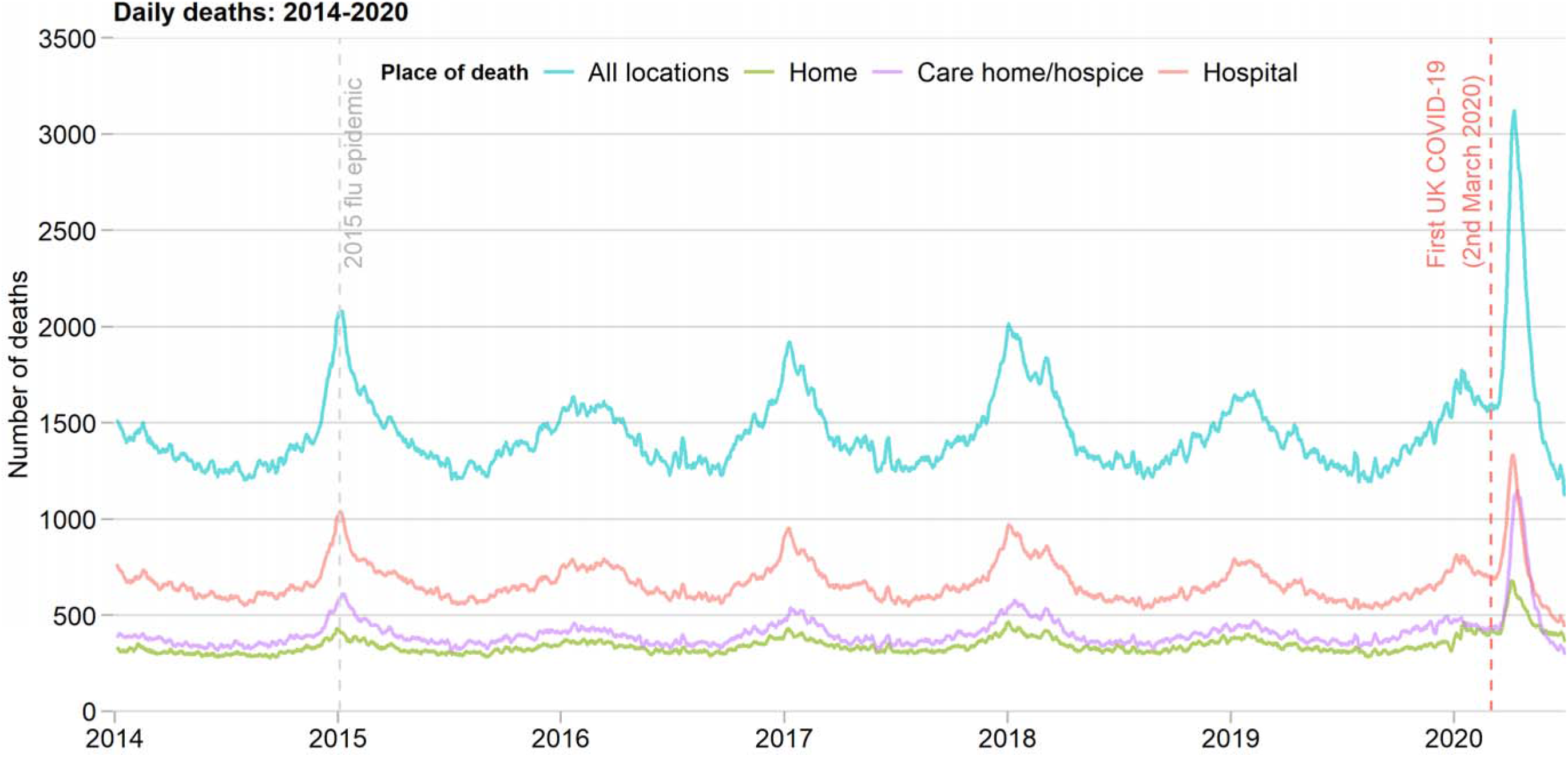
Times series of daily deaths in England and Wales, 2014-2020

**Figure 2.**
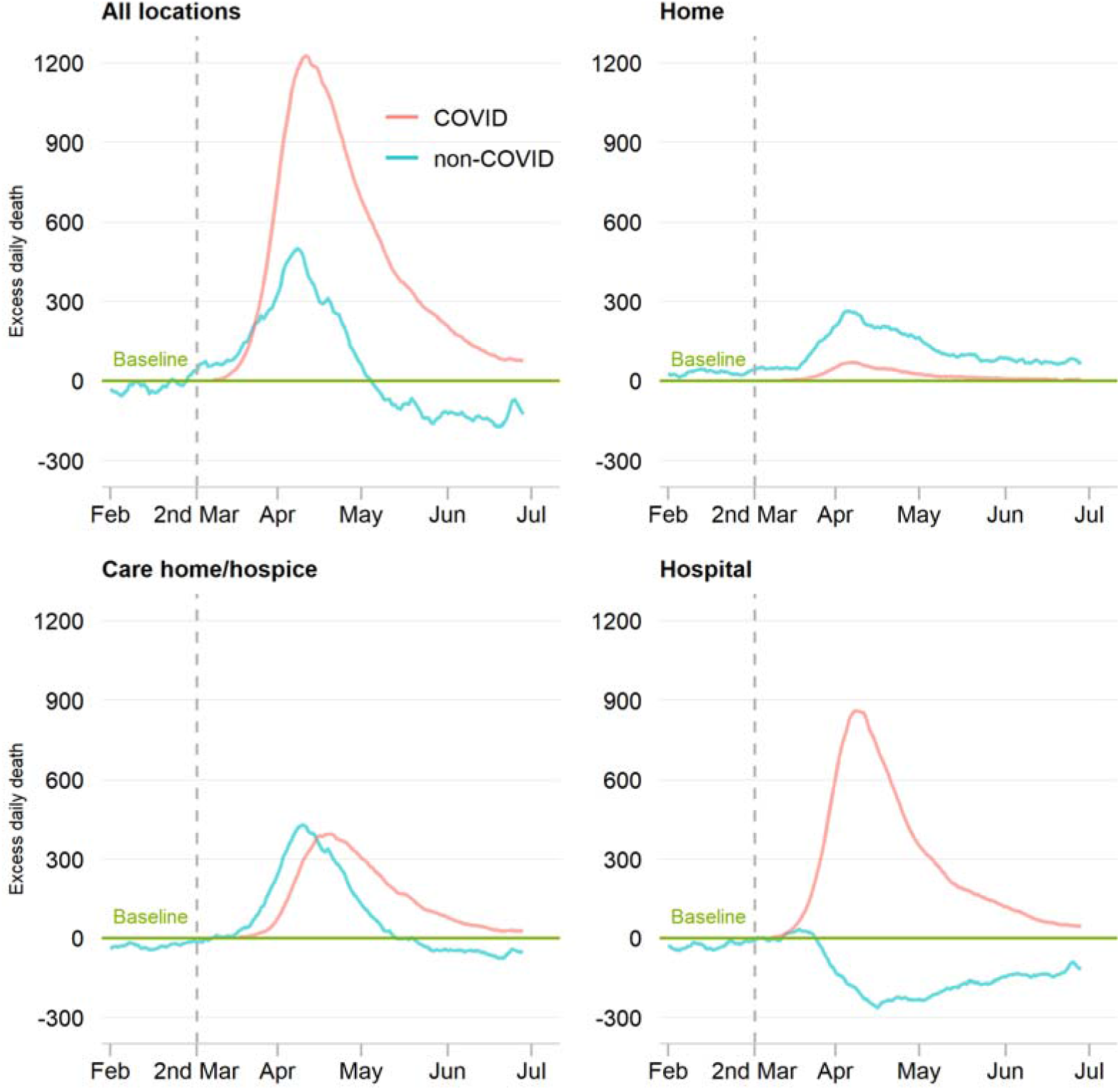
Time series of daily deaths according to COVDI-19, by place of death The number of daily deaths is presented using a 7-day simple moving average (indicating the mean number of daily deaths for that day and the preceding 6 days) from 1st February 2020 up to and including 30th June 2020, adjusted for seasonality. The number of non-COVID-19 excess deaths each day from 1st February 2020 were subtracted from the expected daily death estimated using Farrington surveillance algorithm in the same time period. The green line is a zero historical baseline. The red line represents daily COVID-19 deaths from 2nd March to 30th June 2020, the blue line represents daily non-COVID-19 deaths from 2nd March to 30th June 2020.

**Table 2.** All and COVID-19 related and excess deaths *The numbers for place of death do not add up to the total excess deaths due to missingness.

|  | Excess deaths |  |  |
| --- | --- | --- | --- |
|  | COVID-19 related | Non-COVID-19 related** | Total (% change compared to historical data)*** |
| **Total** | 50,603 | 7,257 | 57,860 (+35%) |
| **Sex** |  |  |  |
| Men | 27,863 | 2,093 | 29,956 (+36%) |
| Women | 22,740 | 5,099 | 27,839 (+33%) |
| **Age category (years)** |  |  |  |
| 18-49 | 995 | -13 | 982 (+15%) |
| 50-59 | 2,312 | 71 | 2,383 (+24%) |
| 60-69 | 4,883 | 368 | 5,251 (+28%) |
| 70-79 | 11,376 | 641 | 12,017 (+31%) |
| 80+ | 31,037 | 6,207 | 37,244 (+40%) |
| **Region** |  |  |  |
| North East | 2,832 | -269 | 2,563 (+27%) |
| North West | 7,741 | -278 | 7,463 (+31%) |
| Yorkshire and The Humber | 4,691 | -81 | 4,610 (+27%) |
| East Midlands | 3,574 | -296 | 3,278 (+23%) |
| West Midlands | 5,848 | 324 | 6,172 (+33%) |
| East of England | 4,902 | 70 | 4,972 (+26%) |
| London | 8,606 | 395 | 9,001 (+55%) |
| South East | 7,124 | -98 | 7,026 (+26%) |
| South West | 2,879 | -230 | 2,649 (+14%) |
| Wales | 2,404 | -688 | 1,716 (+16%) |
| **Place of death\*** |  |  |  |
| Home | 2,334 | 13,856 | 16,190 (+39%) |
| Care home or hospice | 15,966 | 9,645 | 25,611 (+55%) |
| Hospital | 32,112 | -16,174 | 15,938 (+21%) |
| **Underlying cause of deaths** |  |  |  |
| Respiratory diseases | 19,861 | -1,636 | 18,225 (+81%) |
| Dementia and Alzheimer disease | 1,036 | 4,897 | 5,933 (+28%) |
| Symptoms, signs and ill-defined conditions | 2,718 | 1,850 | 4,568 (+101%) |
| Cardiac diseases | 1,050 | 1,175 | 2,225 (+9%) |
| Other cause of diseases | 1,784 | -219 | 1,565 (+6%) |
| Malignant neoplasms | 1,127 | -440 | 687 (+1%) |
| Cerebrovascular diseases | 441 | 530 | 971 (+11%) |
| Diabetes | 164 | 519 | 683 (+32%) |
| Diseases of the urinary system | 280 | 315 | 595 (+23%) |
| Parkinson's disease | 91 | 453 | 544 (+26%) |
| Cirrhosis and other diseases of liver | 116 | 97 | 213 (+8%) |

\*The numbers do not add up to the total deaths due to missingness (1.9%). \*\*Non-COVID-19 related excess deaths were derived by subtracting COVID-19 related excess deaths to total excess deaths. \*\*\* Excess deaths in subgroups may not add up to total excess deaths due to rounding errors when comparing to the historical baseline data.

Among the excess deaths, 50,603 (87%) were COVID-19 related (Figure 2, Table 2). There were large numbers of excess deaths caused by respiratory disease (18,225 deaths; a proportional increase of 81%), dementia (5,933 deaths [including 1,036 related to COVID]; a proportional increase of 28%) as well as ill-defined conditions (4,568 deaths [including 2,718 related to COVID]; a proportional increase of 101%) (Figure 3, Table 2). There were smaller numbers of excess deaths due to cardiac disease (2,225 deaths [including 1,050 related to COVID]; a proportional increase of 9%) and cancer (687 deaths; a proportional increase of 4% [1,127 related to COVID, but 440 fewer cancer deaths than expected after subtracting the COVID related deaths]) (Figure 3, Table 2).

**Figure 3.**
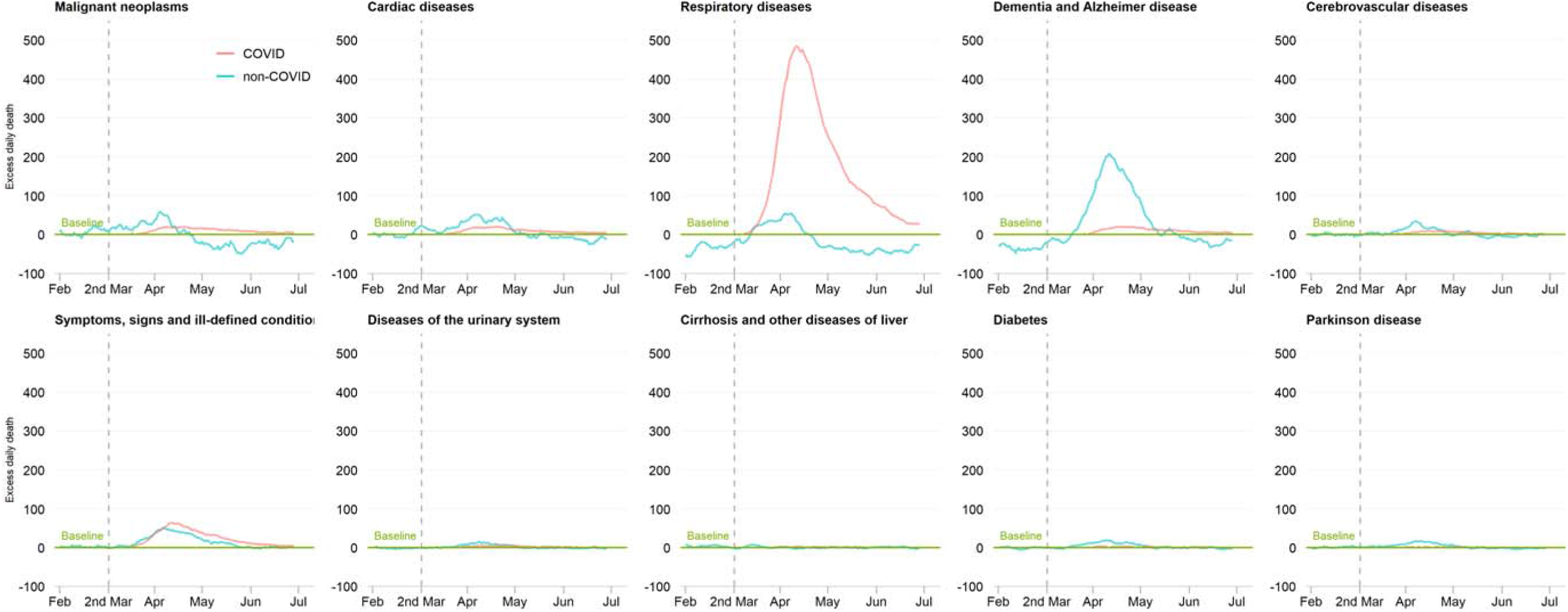
Time series of daily deaths according to COVID-19, by underlying cause of death The number of daily deaths is presented using a 7-day simple moving average (indicating the mean number of daily deaths for that day and the preceding 6 days) from 1st February 2020 up to and including 30th June 2020, adjusted for seasonality. The number of non-COVID-19 excess deaths each day from 1st February 2020 were subtracted from the expected daily death estimated using Farrington surveillance algorithm in the same time period. The green line is a zero historical baseline. The red line represents daily COVID-19 deaths from 2nd March to 30th June 2020, the blue line represents daily non-COVID-19 deaths from 2nd March to 30th June 2020.

### COVID-19 related deaths

Between 2^nd^ March 2020 and the 30^th^ June 2020, there were 50,603 COVID related deaths, one quarter of the deaths occurring during this period (Table 1). About two thirds of the COVID-19 related deaths occurred in hospital, about one third occurred in care homes and hospices, and less than 5% occurred at home (Table 1). Fifty-five percent of COVID-19 related deaths occurred in men and about two-thirds occurred in those aged 80 years or older with less than 2% occurring in those younger than 50 years. In around half of the COVID-19 related deaths, the condition recorded as leading directly to death was recorded as COVID-19 (21,935 deaths) or an ill-defined cause of death (2,718 deaths) with a further 39% of COVID-19 related deaths (19,681 deaths) in which a respiratory disease lead directly to death (639 involving asthma, COPD or another chronic lung disease and 18,264 involving respiratory failure or respiratory infection) (Table 1).Only 1,127 (2%) of the COVID-19 related deaths involved cancer and 1,050 (2%) involved cardiac disease.

### Place and cause of death after 2^nd^ March 2020

Deaths at home increased sharply at the end of March and in early April (Figure 2) and remain above expected levels. Only 14% of the 16,190 excess deaths occurring at home were related to COVID-19 (Table 2). There were 5,963 excess deaths at home due to cancer and 2,485 excess deaths from cardiac disease, very few of which involved COVID-19 (Table 3).

There were 25,611 excess deaths in care homes and hospices, of which about two thirds (15,966 deaths) were related to COVID-19 (Figure 2). Of the excess deaths in care homes and hospices, 5,539 were due to respiratory disease and most of these (4,315 deaths) involved COVID-19. There were 6,267 excess deaths due to dementia, and 2,358 excess deaths from ill-defined conditions, in care homes or hospices, of which only 783 and 1,003 respectively were COVID-19 related. There were 1,495 fewer deaths in care homes and hospices due to cancer than expected and 1,211 excess deaths in care homes due to cardiac disease (Table 3).

**Table 3.**
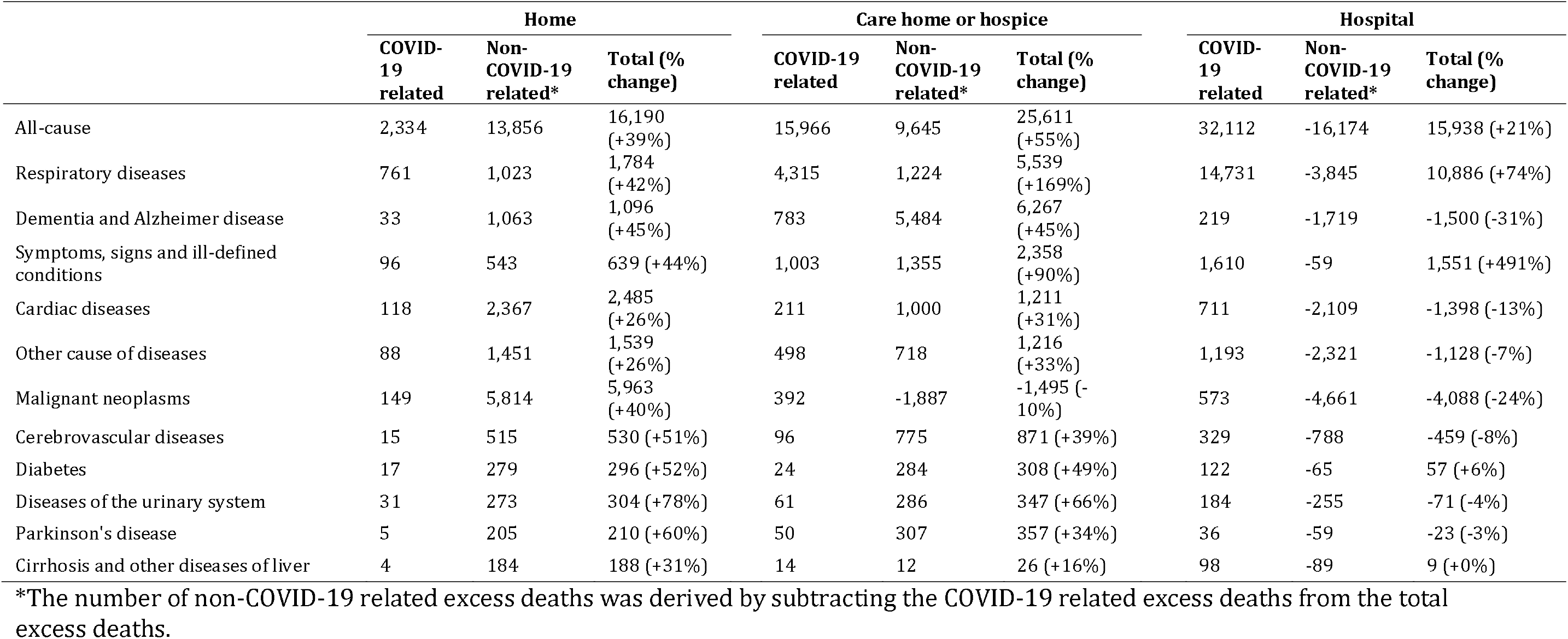
Excess deaths by cause and place of death

|  | Home |  |  | Care home or hospice |  |  | Hospital |  |  |
| --- | --- | --- | --- | --- | --- | --- | --- | --- | --- |
|  | COVID-19 related | Non-COVID-19 related* | Total (% change) | COVID-19 related | Non-COVID-19 related* | Total (% change) | COVID-19 related | Non-COVID-19 related* | Total (% change) |
| All-cause | 2,334 | 13,856 | 16,190 (+39%) | 15,966 | 9,645 | 25,611 (+55%) | 32,112 | -16,174 | 15,938 (+21%) |
| Respiratory diseases | 761 | 1,023 | 1,784 (+42%) | 4,315 | 1,224 | 5,539 (+169%) | 14,731 | -3,845 | 10,886 (+74%) |
| Dementia and Alzheimer disease | 33 | 1,063 | 1,096 (+45%) | 783 | 5,484 | 6,267 (+45%) | 219 | -1,719 | -1,500 (-31%) |
| Symptoms, signs and ill-defined conditions | 96 | 543 | 639 (+44%) | 1,003 | 1,355 | 2,358 (+90%) | 1,610 | -59 | 1,551 (+491%) |
| Cardiac diseases | 118 | 2,367 | 2,485 (+26%) | 211 | 1,000 | 1,211 (+31%) | 711 | -2,109 | -1,398 (-13%) |
| Other cause of diseases | 88 | 1,451 | 1,539 (+26%) | 498 | 718 | 1,216 (+33%) | 1,193 | -2,321 | -1,128 (-7%) |
| Malignant neoplasms | 149 | 5,814 | 5,963 (+40%) | 392 | -1,887 | -1,495 (-10%) | 573 | -4,661 | -4,088 (-24%) |
| Cerebrovascular diseases | 15 | 515 | 530 (+51%) | 96 | 775 | 871 (+39%) | 329 | -788 | -459 (-8%) |
| Diabetes | 17 | 279 | 296 (+52%) | 24 | 284 | 308 (+49%) | 122 | -65 | 57 (+6%) |
| Diseases of the urinary system | 31 | 273 | 304 (+78%) | 61 | 286 | 347 (+66%) | 184 | -255 | -71 (-4%) |
| Parkinson's disease | 5 | 205 | 210 (+60%) | 50 | 307 | 357 (+34%) | 36 | -59 | -23 (-3%) |
| Cirrhosis and other diseases of liver | 4 | 184 | 188 (+31%) | 14 | 12 | 26 (+16%) | 98 | -89 | 9 (+0%) |

\*The number of non-COVID-19 related excess deaths was derived by subtracting the COVID-19 related excess deaths from the total excess deaths.

In hospital after 2 March 2020, there were 32,112 COVID-19 related deaths, but 16,174 fewer deaths than expected which did not involve COVID-19, meaning that the total number of excess deaths in hospital was 15,938 (Figure 2, Table 3). There were 4,088 fewer deaths in hospital due to cancer and 1,398 fewer deaths in hospital due to cardiac disease than expected (Table 3).

The time course of changes in deaths by cause and place is shown in supplementary figure 1. At the end of March, there was an increase in deaths from cancer in homes, with a corresponding fall in cancer deaths below the historical baseline in hospitals (Supplement Figure 1). These changes in the location of cancer deaths were still present at the end of June. During March and April, deaths from cardiac disease increased in homes, care homes and hospices and fell below the historical baseline in hospitals, returning to expected levels in all locations by the end of June. In care homes and hospices, there was a sharp increase at the end of March in deaths from dementia and Alzheimer’s disease and from respiratory disease, with a subsequent rapid decline in late April.

### Comparison with seasonal influenza epidemics

By comparison, the numbers of excess deaths from influenza and pneumonia in previous years occurred with the greatest magnitude in hospital and were driven equally by respiratory and non-respiratory causes (Supplement Figure 2).

## Discussion

We report, for the first time, in a complete analysis of all adult deaths in England and Wales, the extent, site and underlying causes of the increased mortality during the COVID-19 pandemic compared with previous years. This shows that, most of the 58,000 excess deaths during this period involved COVID-19 and, in most of these, COVID-19 appeared to be the direct cause of death. However, there was a substantial increase in the absolute numbers of deaths occurring at home, especially from cancer and cardiac disease, while deaths from these causes in hospital were lower than expected. In care homes and hospices there was a substantial increase in the absolute numbers of deaths from dementia, Alzheimer’s disease and ill-defined causes, in addition to COVID-19 related deaths.

We found evidence for the displacement in the place of death from hospital to community setting during the pandemic. Historically in England and Wales, almost half of all adult deaths occur in hospital, but during the pandemic only a quarter did. During the pandemic about 26,000 excess deaths (almost half of the total excess deaths) occurred in care homes and hospices. Residents of care homes frequently died from respiratory disease (mostly involving COVID-19), but also from ‘symptoms and signs of ill-defined conditions’ (which typically indicates old age and frailty^18^) and dementia and Alzheimer’s disease. While it is not possible to be certain about the factors leading to the substantial excess in deaths from these less well-defined causes, undiagnosed COVID-19 is likely.

The efficient person-to-person transmission of the SARS-2-CoV virus,^19^ its asymptomatic incubation and transmission period^20 21^ and propensity to death in the elderly and co-morbid will have been major contributing factors to the excess mortality in care homes. In March 2020, a report detailing an outbreak of COVID-19 infection at a long-term care centre that was associated with high mortality rates recommended proactive steps by such places to identify and exclude potentially infected staff and visitors and implement infection prevention and control measures to reduce the introduction of the virus to residents.^22^ Yet, in the UK, patients were discharged from hospitals to care homes without information of their infective status, where the virus could easily spread ^12^ and actions to reduce the spread of the virus in social care were not implemented early in the pandemic.^23^ Early in the pandemic testing of suspected cases was only available in hospital while routine testing of staff and residents in care homes was not implemented until May 2020^24^ meaning potentially leading to under-diagnosis of COVID-19.^25^ Additionally, it is possible that care home residents who became unwell during the pandemic were not referred or decided not to go to hospital for fear of becoming infected – a notion which aligns to the substantial reduction in hospital attendances for medical emergencies following the UK lockdown.^6–10^

Most of the deaths in hospital involved COVID-19. After excluding the COVID-19 related deaths, there were fewer deaths than expected for cancer, cardiac disease, cerebrovascular disease, and dementia and Alzheimer’s disease in hospital. This finding supports the concept of patients with non-COVID-19 illness staying in the community rather than attending hospital. Other possible explanations include factors related to the re-deployment of front-line staff and the cancellation of procedures, undermining routine care and the use of alternative levels of treatment, as well as earlier discharge back to the community.

At home, the largest number of excess deaths were from cardiac disease and cancer and few deaths involved COVID-19. This may be explained by infection serving as a trigger to acute decompensation of a pre-existing disease^26^ (and which may be under-reported due to non-systematic testing^27^) but is more likely to be related to a reluctance by the public to attend hospital when unwell because of fear of becoming infected with the SARS-CoV-2 virus. Another possible explanation is that early in the course of the pandemic hospitals prepared for a potential mass influx of patients by expeditious hospital discharge of in-patients to the community, which may have resulted in a number of deaths.

Whilst previous reports have described an elevated risk of death among the elderly and people with cardiovascular disease during the COVID-19 pandemic, none have characterised the underlying causes and place of death in an unselected national cohort.^5 28 29^ The unique strengths of this investigation include full population coverage of all adult deaths across all places of death. Nonetheless, our study has limitations. First, during the COVID19 pandemic, emergency guidance enabled any doctor in the UK (not just the attending) to complete the MCCD, the duration of time over which the deceased was not seen before referral to the coroner was extended from 14 to 28 days, and causes of death could be “to the best of their knowledge and belief” without diagnostic proof, if appropriate and to avoid delay.^30^ This may have resulted in inaccurate recording of cause of death. Second, this analysis will have excluded a small proportion of deaths under review by the Coroner, though typically these will have been unnatural in aetiology. Third, we did not have access to laboratory testing data. It is estimated that methods identifying COVID-19 deaths using laboratory-testing data have identified about an additional 1500 deaths during the pandemic period compared to death certificate methods. However, this would only explain a tiny proportion of the non-COVID-19 excess deaths observed in this study.^31^

The COVID-19 pandemic has resulted in major global changes to society and health. These analyses raise important findings for government and the NHS. There was a huge burden of excess deaths occurring in care homes, which were poorly characterised, and were likely to be, at least in part, the result of undiagnosed COVID-19. Effective assessment and testing, along with adequate staffing and infection control measures, in care homes should be a priority in the event of a second rise in cases. Second, there was a smaller but important and ongoing excess in deaths at home, particularly from cancer and cardiac disease, which suggests avoidance of hospital care for non-COVID-19 conditions. Clear public messaging encouraging patients to seek medical advice when necessary, along with a robust strategy to maintain COVID-19 free areas within hospitals, are needed.

### Data sharing

The Secretary of State for Health and Social Care has issued a time limited Notice under Regulation 3(4) of the NHS (Control of Patient Information Regulations) 2002 (COPI) to share confidential patient information. The co-authors are not permitted to share the data.

### Patient and Public Involvement statement

Patient and public were not involved because this study was to analyse routinely collected mortality data.

Updates of all Figures are available at cardiovascularcovid.leeds.ac.uk

## Data Availability

The data will be available upon request pending approval by NHS digital, UK.

---

**Supplement**

**Supplement Table 1.** ICD-10 codes for cause of death groups

| **Cause of death groups** | **ICD-10 codes** |
| --- | --- |
| Accidental drowning and submersion | W65–W74 |
| Accidental falls | W00–W19 |
| Accidental poisoning | X40–X49 |
| Accidental threats to breathing | W75–W84 |
| Aortic aneurysm and dissection | I71 |
| Appendicitis, hernia and intestinal obstruction | K35–K46, K56 |
| Atherosclerosis | I70 |
| Cardiac diseases | I10–I15, I20–I25, I42, I46–I51 |
| Cerebral palsy and other paralytic syndromes | G80–G83 |
| Cerebrovascular diseases | I60–I69 |
| Certain conditions originating in the perinatal period | P00–P96 |
| Chronic rheumatic heart diseases | I05–I09 |
| Cirrhosis and other diseases of liver | K70–K76 |
| Congenital malformations, deformations and chromosomal abnormalities | Q00–Q99 |
| Dementia and Alzheimer disease | F01, F03, G30 |
| Diabetes | E10–E14 |
| Diseases of the musculoskeletal system | M00–M99 |
| Diseases of the urinary system | N00–N39 |
| Disorders of fluid, electrolyte and acid–base balance (incl. dehydration) | E86–E87 |
| Epilepsy and status epilepticus | G40–G41 |
| Homicide and probable homicide | U50.9, X85–Y09, Y87.1 |
| Human immunodeficiency virus [HIV] disease | B20–B24 |
| In situ and benign neoplasms, and neoplasms of uncertain or unknown behaviour | D00–D48 |
| Intestinal infectious diseases | A00–A09 |
| Influenza and pneumonia | J09–J18 |
| Land transport accidents | V01–V89 |
| Malignant neoplasms | C00–C97 |
| Malnutrition, nutritional anaemias and other nutritional deficiencies | D50–D53, E40–E64 |
| Meningitis and meningococcal infection | A39, A87, G00–G03 |
| Mental and behavioural disorders due to psychoactive substance use | F10–F19 |
| Non-intentional firearm discharge | W32–W34 |
| Nonrheumatic valve disorders and endocarditis | I34–I38 |
| Parkinson disease | G20 |
| Pregnancy, childbirth and the puerperium | O00–O99 |
| Pulmonary heart disease and diseases of pulmonary circulation | I26–I28 |
| Respiratory diseases | J00–J96 |
| Septicaemia | A40–A41 |
| Suicide and injury/poisoning of undetermined intent<sup>2</sup> | X60–X84, Y10–Y34 |
| Symptoms, signs and ill-defined conditions | R00–R99 |
| Systemic atrophies primarily affecting the central nervous system | G10–G12 |
| Tuberculosis | A15–A19, B90 |
| Vaccine-preventable diseases<sup>1</sup> | A33–A37, A49.2, A80, B01, B02, B05, B06, B15, B16, B17.0, B18.0, B18.1, B26, B91, G14 |
| Vector-borne diseases and rabies | A20, A44, A75–A79, A82–A84, A85.2, A90–A98, B50–B57 |

**Supplement Table 2.**
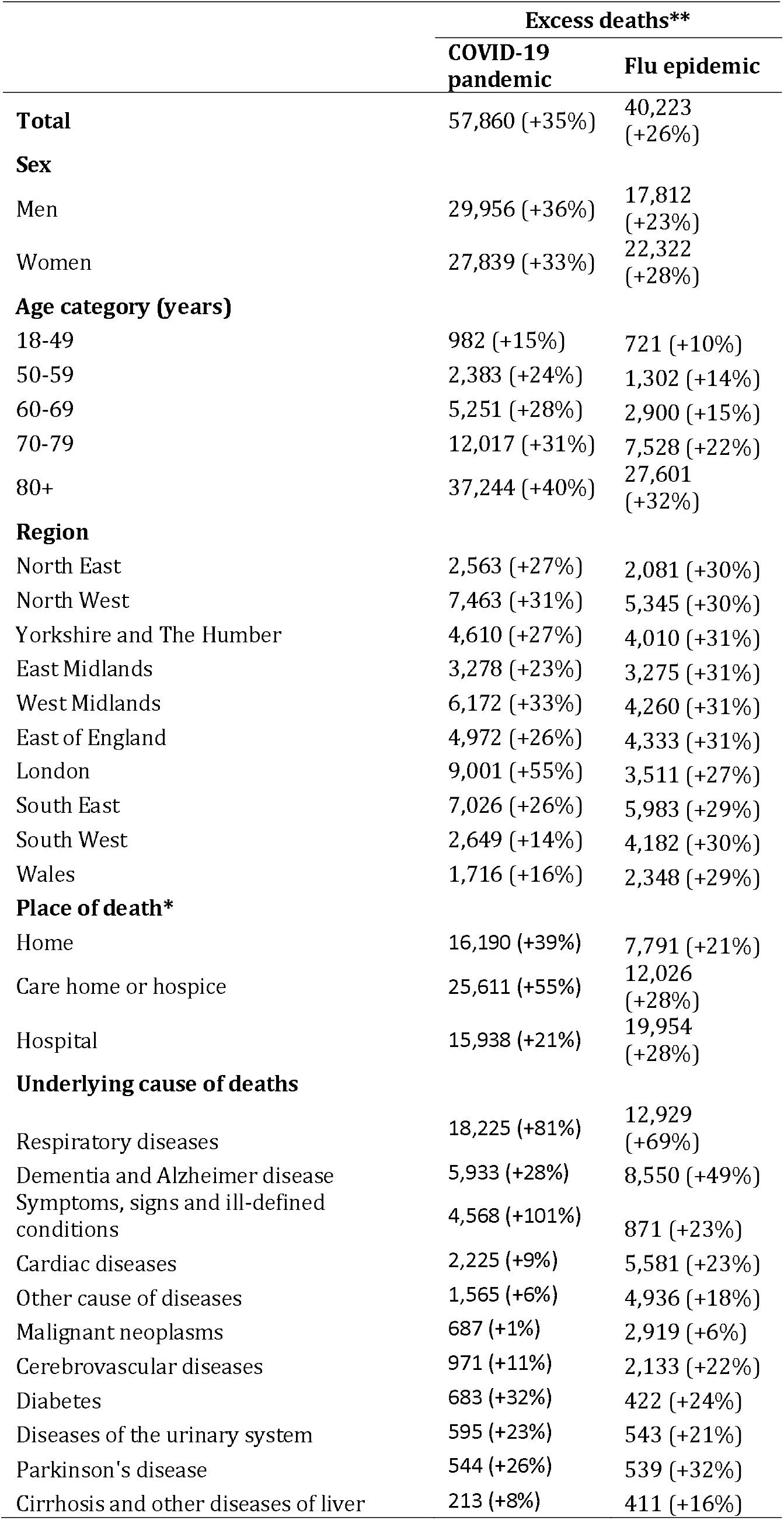
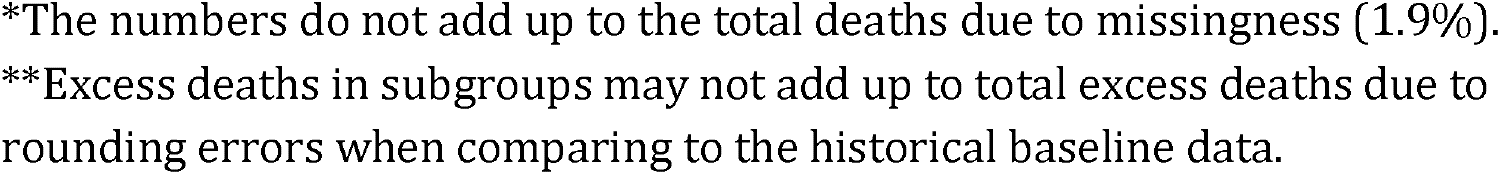
Excess deaths for COVID-19 period and flu epidemic

|  | Excess deaths** |  |
| --- | --- | --- |
|  | COVID-19 pandemic | Flu epidemic |
| **Total** | 57,860 (+35%) | 40,223 (+26%) |
| **Sex** |  |  |
| Men | 29,956 (+36%) | 17,812 (+23%) |
| Women | 27,839 (+33%) | 22,322 (+28%) |
| **Age category (years)** |  |  |
| 18-49 | 982 (+15%) | 721 (+10%) |
| 50-59 | 2,383 (+24%) | 1,302 (+14%) |
| 60-69 | 5,251 (+28%) | 2,900 (+15%) |
| 70-79 | 12,017 (+31%) | 7,528 (+22%) |
| 80+ | 37,244 (+40%) | 27,601 (+32%) |
| **Region** |  |  |
| North East | 2,563 (+27%) | 2,081 (+30%) |
| North West | 7,463 (+31%) | 5,345 (+30%) |
| Yorkshire and The Humber | 4,610 (+27%) | 4,010 (+31%) |
| East Midlands | 3,278 (+23%) | 3,275 (+31%) |
| West Midlands | 6,172 (+33%) | 4,260 (+31%) |
| East of England | 4,972 (+26%) | 4,333 (+31%) |
| London | 9,001 (+55%) | 3,511 (+27%) |
| South East | 7,026 (+26%) | 5,983 (+29%) |
| South West | 2,649 (+14%) | 4,182 (+30%) |
| Wales | 1,716 (+16%) | 2,348 (+29%) |
| **Place of death\*** |  |  |
| Home | 16,190 (+39%) | 7,791 (+21%) |
| Care home or hospice | 25,611 (+55%) | 12,026 (+28%) |
| Hospital | 15,938 (+21%) | 19,954 (+28%) |
| **Underlying cause of deaths** |  |  |
| Respiratory diseases | 18,225 (+81%) | 12,929 (+69%) |
| Dementia and Alzheimer disease | 5,933 (+28%) | 8,550 (+49%) |
| Symptoms, signs and ill-defined conditions | 4,568 (+101%) | 871 (+23%) |
| Cardiac diseases | 2,225 (+9%) | 5,581 (+23%) |
| Other cause of diseases | 1,565 (+6%) | 4,936 (+18%) |
| Malignant neoplasms | 687 (+1%) | 2,919 (+6%) |
| Cerebrovascular diseases | 971 (+11%) | 2,133 (+22%) |
| Diabetes | 683 (+32%) | 422 (+24%) |
| Diseases of the urinary system | 595 (+23%) | 543 (+21%) |
| Parkinson's disease | 544 (+26%) | 539 (+32%) |
| Cirrhosis and other diseases of liver | 213 (+8%) | 411 (+16%) |

\*The numbers do not add up to the total deaths due to missingness (1.9%). \*\*Excess deaths in subgroups may not add up to total excess deaths due to rounding errors when comparing to the historical baseline data.

**Supplement Figure 1.**
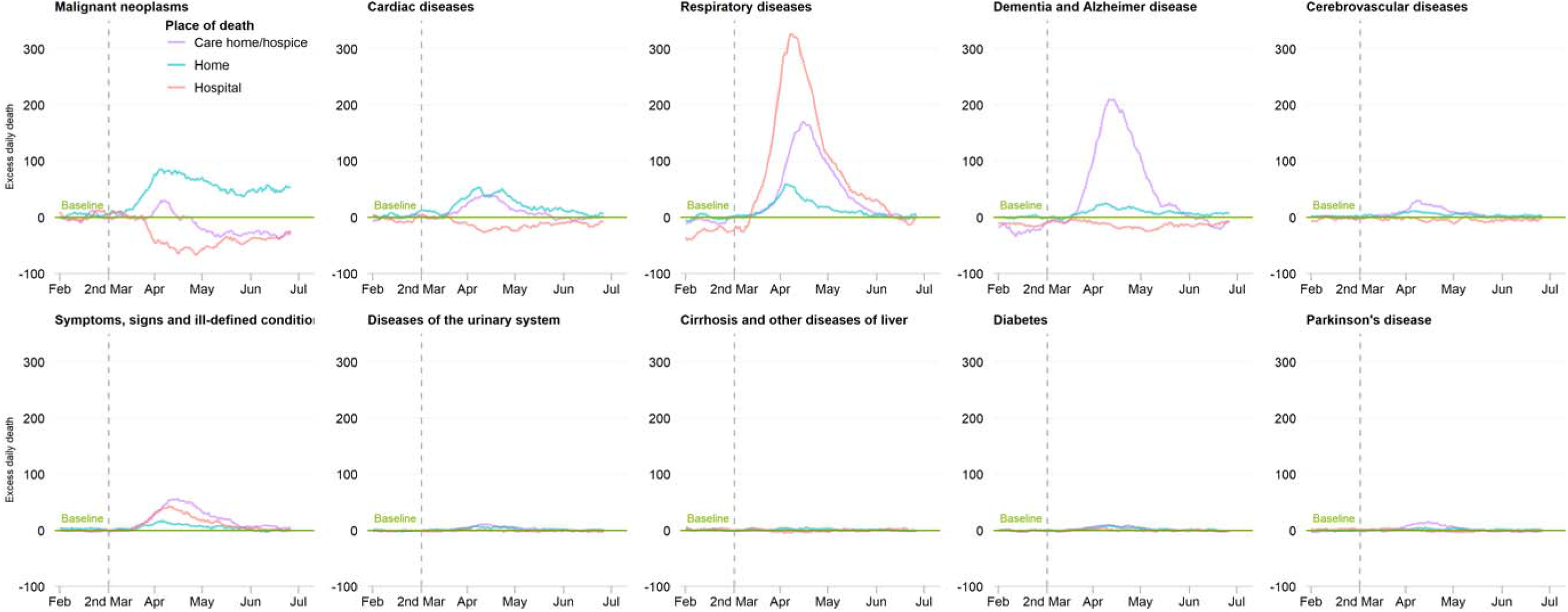
Time series of daily deaths by underlying cause of death and place of death.

**Supplement Figure 2.**
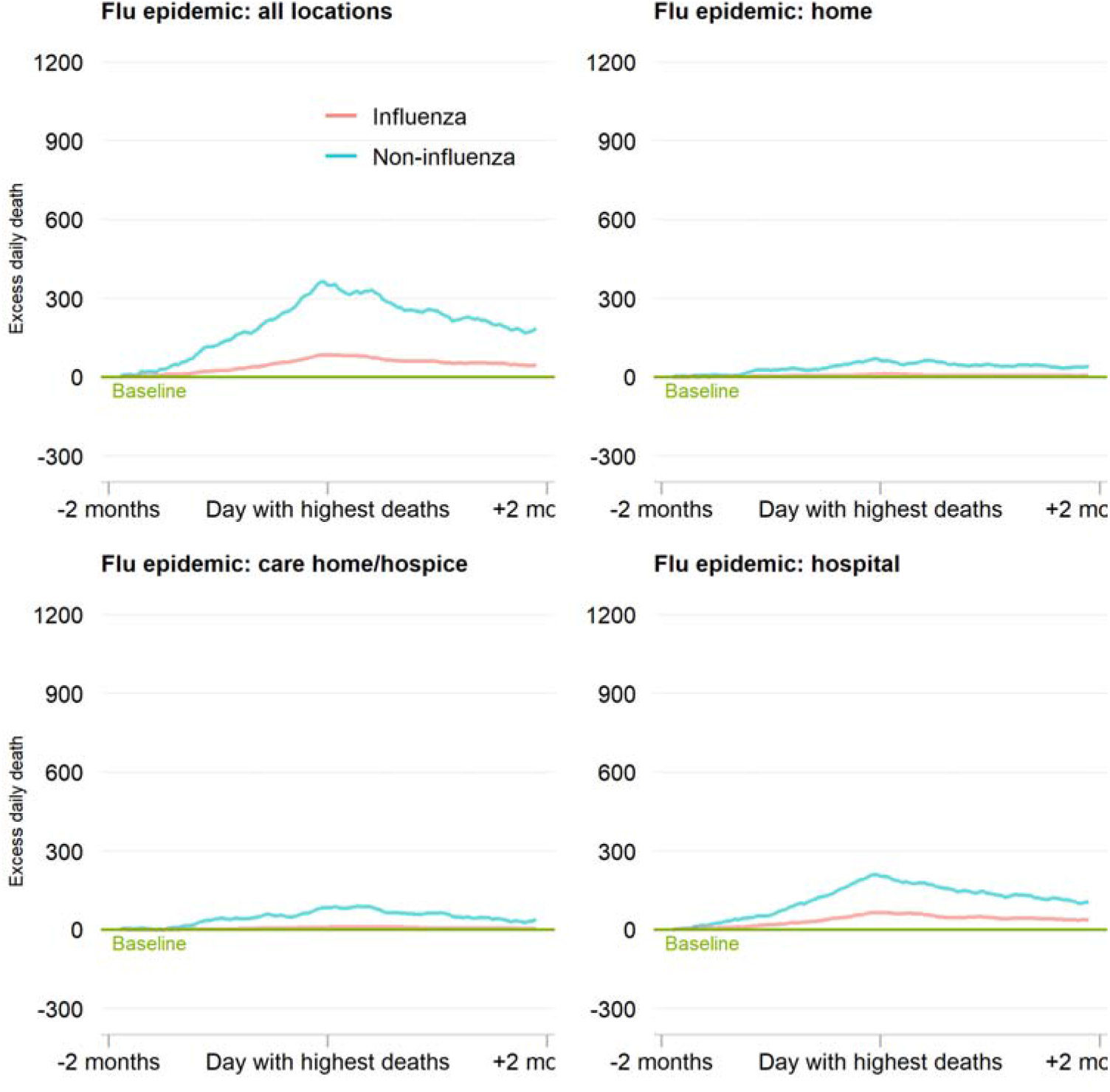
Time series of daily deaths according to flu epidemic, by place of death. Note: the scale of y-axis was kept the same as main Fig 2

## References

1. WHO coronavirus disease (COVID-19) dashboard. Geneva: World Health Organization, 2020. Available online: https://covid19.who.int/ (accessed 06th Aug, 2020).

2. https://ig.ft.com/coronavirus-chart/?areas=usa&areas=gbr&areas=bra&areasRegional=usny&areasRegional=usca&areasRegional=usfl&areasRegional=ustx&cumulative=0&logScale=1&perMillion=0&values=deaths Accessed 6th Aug 2020.

3. Deaths involving COVID-19, England and Wales: deaths occurring in May 2020. https://www.ons.gov.uk/peoplepopulationandcommunity/birthsdeathsandmarriages/deaths/bulletins/deathsinvolvingcovid19englandandwales/latest.

4. Zhou F, Yu T, Du R, et al. Clinical course and risk factors for mortality of adult inpatients with COVID-19 in Wuhan, China: a retrospective cohort study. Lancet 2020;395(10229): 1054-62. doi: 10.1016/S0140-6736(20)30566-3 [published Online First: 2020/03/15]

5. Banerjee A, Pasea L, Harris S, et al. Estimating excess 1-year mortality associated with the COVID-19 pandemic according to underlying conditions and age: a population-based cohort study. Lancet 2020 doi: 10.1016/S0140-6736(20)30854-0 [published Online First: 2020/05/15]

6. Solomon MD, McNulty EJ, Rana JS, et al. The Covid-19 Pandemic and the Incidence of Acute Myocardial Infarction. N Engl J Med 2020 doi: 10.1056/NEJMc2015630 [published Online First: 2020/05/20]

7. https://assets.publishing.service.gov.uk/government/uploads/system/uploads/attachment_data/file/886455/EDSSSBulletin2020wk20.pdf.

8. Kansagra AP, Goyal MS, Hamilton S, et al. Collateral Effect of Covid-19 on Stroke Evaluation in the United States. N Engl J Med 2020 doi: 10.1056/NEJMc2014816 [published Online First: 2020/05/10]

9. Bollmann A, Hohenstein S, Meier-Hellmann A, et al. Emergency hospital admissions and interventional treatments for heart failure and cardiac arrhythmias in Germany during the Covid-19 outbreak Insights from the German-wide Helios hospital network. Eur Heart J Qual Care Clin Outcomes 2020 doi: 10.1093/ehjqcco/qcaa049 [published Online First: 2020/06/06]

10. Mafham MM, Spata E, Goldacre R, et al. COVID-19 pandemic and admission rates for and management of acute coronary syndromes in England. Lancet 2020 doi: 10.1016/S0140-6736(20)31356-8 [published Online First: 2020/07/18]

11. Wu J, Mamas M, Rashid M, et al. Patient response, treatments and mortality for acute myocardial infarction during the COVID-19 pandemic. European Heart Journal - Quality of Care and Clinical Outcomes 2020 doi: 10.1093/ehjqcco/qcaa062

12. Oliver D. David Oliver: Let’s be open and honest about covid-19 deaths in care homes. BMJ 2020;369:m2334. doi: 10.1136/bmj.m2334 [published Online First: 2020/06/20]

13. Deaths registered weekly in England and Wales, provisional: week ending 12 June 2020. https://www.ons.gov.uk/peoplepopulationandcommunity/birthsdeathsandmarriages/deaths/bulletins/deathsregisteredweeklyinenglandandwalesprovisional/latest#deaths-registered-by-place-of-occurrence.

14. Comas-Herrera AZ, J; Litwin, C; Hsu, AT; Lane, N; Fernández, J. Mortality Associated with COVID-19 Outbreaks in Care Homes: Early International Evidence. https://ltccovid.org/wp-content/uploads/2020/04/Mortality-associated-with-COVID-26-April-1.pdf.

15. Oliver D. David Oliver: Preventing more deaths in care homes in a second pandemic surge. BMJ 2020;369:m2461. doi: 10.1136/bmj.m2461 [published Online First: 2020/06/25]

16. https://www.ons.gov.uk/peoplepopulationandcommunity/birthsdeathsandmarriages/deaths/methodologies/userguidetomortalitystatisticsjuly2017.

17. User guide to mortality statistics. https://www.ons.gov.uk/peoplepopulationandcommunity/birthsdeathsandmarriages/deaths/methodologies/userguidetomortalitystatisticsjuly2017#ons-short-list-of-cause-of-death.

18. Noufaily A, Enki DG, Farrington P, et al. An improved algorithm for outbreak detection in multiple surveillance systems. Stat Med 2013;32(7):1206-22. doi: 10.1002/sim.5595 [published Online First: 2012/09/04]

19. Chen Y, Li L. SARS-CoV-2: virus dynamics and host response. Lancet Infect Dis 2020;20(5):515-16. doi: 10.1016/S1473-3099(20)30235-8 [published Online First: 2020/03/28]

20. Arons MM, Hatfield KM, Reddy SC, et al. Presymptomatic SARS-CoV-2 Infections and Transmission in a Skilled Nursing Facility. N Engl J Med 2020;382(22):2081-90. doi: 10.1056/NEJMoa2008457 [published Online First: 2020/04/25]

21. Lavezzo E, Franchin E, Ciavarella C, et al. Suppression of a SARS-CoV-2 outbreak in the Italian municipality of Vo’. Nature 2020 doi: 10.1038/s41586-020-2488-1 [published Online First: 2020/07/01]

22. McMichael TM, Currie DW, Clark S, et al. Epidemiology of Covid-19 in a Long-Term Care Facility in King County, Washington. N Engl J Med 2020;382(21):2005-11. doi: 10.1056/NEJMoa2005412 [published Online First: 2020/03/29]

23. Griffin S. Covid-19: “Staggering number” of extra deaths in community is not explained by covid-19. BMJ 2020;369:m1931. doi: 10.1136/bmj.m1931 [published Online First: 2020/05/15]

24. https://www.gov.uk/government/publications/coronavirus-covid-19-support-for-care-homes/coronavirus-covid-19-care-home-support-package.

25. Raleigh VS. Tackling UK’s mortality problem: covid-19 and other causes. BMJ 2020;369:m2295. doi: 10.1136/bmj.m2295 [published Online First: 2020/06/13]

26. Zheng YY, Ma YT, Zhang JY, et al. COVID-19 and the cardiovascular system. Nat Rev Cardiol 2020;17(5):259-60. doi: 10.1038/s41569-020-0360-5 [published Online First: 2020/03/07]

27. Iacobucci G. Covid-19: Lack of capacity led to halting of community testing in March, admits deputy chief medical officer. BMJ 2020;369:m1845. doi: 10.1136/bmj.m1845 [published Online First: 2020/05/08]

28. https://www.ons.gov.uk/peoplepopulationandcommunity/birthsdeathsandmarriages/deaths/bulletins/deathsinvolvingcovid19englandandwales/deathsoccurringinapril2020#pre-existing-conditions-of-people-who-died-with-covid-19

29. Docherty AB, Harrison EM, Green CA, et al. Features of 20 133 UK patients in hospital with covid-19 using the ISARIC WHO Clinical Characterisation Protocol: prospective observational cohort study. BMJ 2020;369:m1985. doi: 10.1136/bmj.m1985 [published Online First: 2020/05/24]

30. https://assets.publishing.service.gov.uk/government/uploads/system/uploads/attachment_data/file/877302/guidance-for-doctors-completing-medical-certificates-of-cause-of-death-covid-19.pdf.

31. https://www.cebm.net/covid-19/covid-19-deaths-in-england-and-wales-resolving-discrepancies-in-deaths-outside-of-hospital/.

